# The association of NLRP3 polymorphisms and its downstream interaction in mild cognitive impairment

**DOI:** 10.1101/2025.03.21.25324400

**Authors:** Ruonan Gao, Linda Chiu Wa Lam, Suk Ling Ma

**Affiliations:** Department of Psychiatry, The Chinese University of Hong Kong, Hong Kong SAR, China

**Author notes:** Corresponding author: Dr Suk Ling Ma, Department of Psychiatry, The Chinese University of Hong Kong, Hong Kong SAR, China.

## Abstract

**Background:** Nucleotide-binding domain and leucine-rich repeat (LRR)-containing family protein 3 (NLRP3) has been widely studied in the pathogenesis of mild cognitive impairments (MCI) and Alzheimer’s Disease (AD). Single nucleotide polymorphisms (SNPs) of NLRP3 gene are associated with various diseases, however the association between NLRP3 SNPs and downstream pathway is unclear.

**Methods:** 12 intron SNPs and 2 exon SNPs were genotyped in 235 normal controls and 331 MCI older adults. NLRP3 and other inflammation-related genes expression were quantified in peripheral blood mononuclear cells (PBMC) from the older adults by quantitative PCR (qPCR). Functional studies of selected mutations were performed by luciferase assay. The older adults were followed up for 2 years to investigate the relationship between NLRP3 SNPs and risk of cognitive decline.

**Results:** Our study showed rs10754558 and rs7525979 were associated with an increased risk of MCI. The T allele of rs12564791 was associated with higher gene expression level of NLRP3, interleukin-18 (IL-18), PYCARD, and CASP1. rs12048215, rs10754555, and rs7525979 were associated with cognitive decline as shown by the reduction of Montreal Cognitive Assessment (MoCA) score. Functional studies showed that both rs10754558 and rs10754555 G to C mutation affected enhancer activity of transcription. rs10754558 G to C mutation also disturbed the interaction between NLRP3 3’UTR and miR-425-5p. Plasma miR-425-5p expression was negatively correlated with MoCA score.

**Conclusions:** Our study suggested that genetic variations of NLRP3 were associated with the cognitive decline by affecting the gene expression of inflammation-related genes and its interactions to miRNAs.

## Introduction

Alzheimer’s Disease (AD) is a progressive neurodegenerative disease and it is the most common cause of dementia. AD is characterized by the accumulation of extracellular amyloid-beta (Aβ), intracellular neurofibrillary tangles consisting of hyperphosphorylated tau and neuronal loss. The major symptoms of AD include memory loss, cognitive impairment and decline in mobility which affect both patients and their family. There is currently no treatment for full recovery from AD and only symptomatic treatments are available. Mild cognitive impairment (MCI) is the preclinical stage of AD. People with MCI have higher risk of developing AD. Therefore, early detection and intervention is important for slowing down the disease progression.

Inflammasome is a multi-protein complex that regulates inflammation and plays an important role in the innate immune system. The NOD-like receptor pyrin domain-containing 3 (NLRP3) is the best studied inflammasome. NLRP3 consists of the sensor protein NLRP3, the adaptor protein apoptosis-associated speck-like protein containing a caspase recruitment domain (PYCARD) and the effector protein pro-caspase1. The activation of NLRP3 leads to the production and release of inflammatory cytokines interleukin-18 (IL-18) and interleukin-1 beta (IL-1β). NIMA-related kinase 7 (NEK7) is a NLRP3-binding protein that have been identified as a modulator of NLRP3 oligomerization and activation recently(1). Several studies have demonstrated that activation of NLRP3 contributes to AD pathogenesis. Gene expression of NLRP3 and downstream cytokines, IL<u>-</u>β and IL-18 are up-regulated in peripheral blood mononuclear cells (PBMC) of both mild and severe AD patients(2). Increased expression are also detected in cerebral temporal cortex of AD patients(3). In APP/PS1 mouse model, NLRP3 knockout or NLRP3 inhibition via small molecules lead to decreased Aβ accumulation, increased Aβ clearance and better cognitive function(4,5). Inhibition of NLRP3 reduced the production of tumor necrosis factor-alpha (TNF-α) and interferon-gamma (IFN-γ) in vivo, which are involved in neuronal necroptosis and neuronal death in AD(6–9). These results suggest that NLRP3 participates in AD development and might be a therapeutic target.

Human NLRP3 gene is located on chromosome 1q44, including 9 exons. Up to now, over 60 single nucleotide polymorphisms (SNPs) have been identified in NLRP3 gene(10). The 3’UTR SNP rs10754558 affects NLRP3 gene expression and mRNA stability as well as its interaction with miR-146a(11–13). The exon SNP rs35829419 influences IL-1β production in plasma(14). Intron SNPs rs2027432 and rs4612666 can enhance the promoter activity of NLRP3 gene(11,15). NLRP3 SNPs have been associated with various diseases, including type 2 diabetes mellitus(10), rheumatoid arthritis(16), ischemic stroke(17). Previous study also showed that NLRP3 SNPs were associated with late-onset AD (LOAD)(18). However, whether NLRP3 SNPs are correlated to MCI susceptibility in Chinese is not clear.

This study investigated the relationship between NLRP3 SNPs and the risk of developing MCI in Southern Chinese. The impact of SNPs on cognitive decline and the effect of the SNPs on NLRP3 related gene expression were also determined. Functional study were performed on some of the significant SNPs to elucidate the effect of these SNPs on NLRP3 transcription and interaction with miRNA. Our results suggested that NLRP3 SNPs may affect the risk of developing MCI in Chinese.

## Material and methods

### Participants

331 Chinese MCI older adults (41.3% men, mean age 70.49 years old, SD=6.74, range 60-92 years) and 235 Chinese normal controls (40.9% men, mean age 70.50 years old, SD=6.75, range 60-93 years) were recruited from Hong Kong. The inclusion criteria for all participants were aged 60-95, ethnic Chinese, living in the community, with no dementia (Clinical Dementia Rating (CDR) 0 or 0.5) and free of depressive symptoms and no neurological conditions that may affect cognition. CDR was used to define MCI (CDR=0.5) or normal controls (CDR=0). During recruitment, our research staff explained the procedure and obtained informed consent from the participants. The study was approved by the Clinical Research Ethics Committee of the Chinese University of Hong Kong.

### Assessments

All participants underwent clinical and cognitive assessments at baseline interview. Including CDR and Hong Kong Montreal Cognitive Assessment (HKMOCA)(19). CDR is a standard clinical assessment for evaluating the overall severity of cognitive impairment. HKMOCA is a locally validated cognitive screening test sensitive in detecting early memory and executive deficits.

The total score of HKMOCA is 30, the higher the score, the better the cognition. A subset of participants received baseline and 2-year follow-up assessments to compare the cognitive performance over 2 years.

### SNPs selection

Tag SNPs were selected according to HapMap data of Chinese Han population (CHB)(20). 11 tag SNPs with minor allele frequency (MAF) higher than 0.1 were chosen for genotyping. Three SNPs reported to be associated with AD, namely rs2027432, rs10754558 and rs35829419 were also included(18). The position of the SNPs was shown in Figure 1.

**Figure 1.**
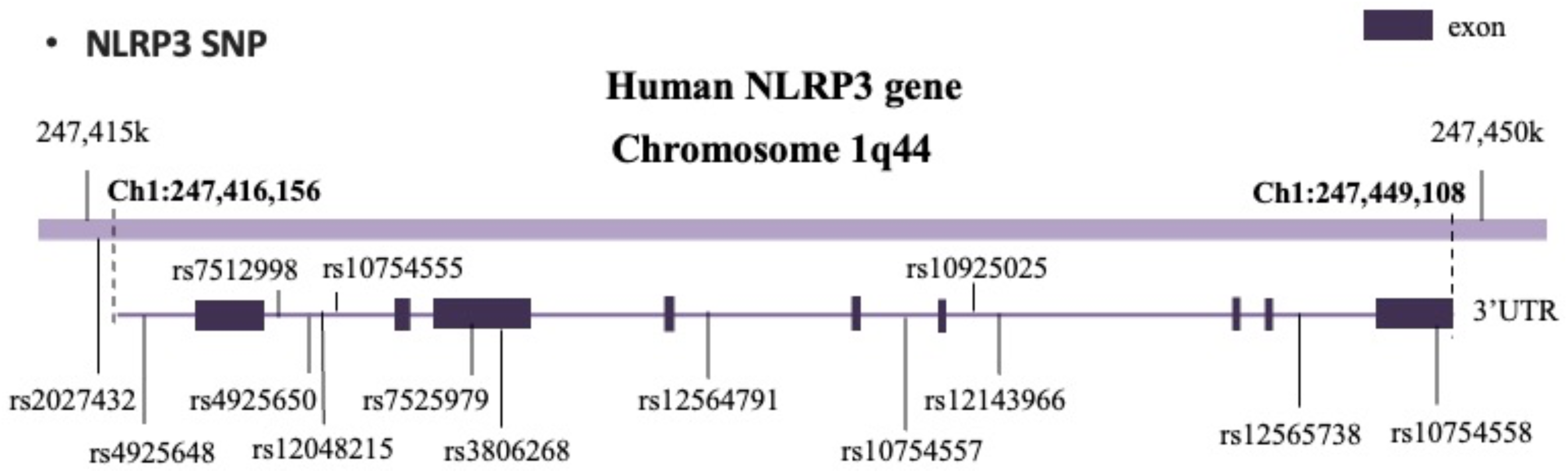
The location of NLRP3 SNP

### Genotyping

Genomic DNA extraction was performed by PureLink Genomic DNA Kit (Invitrogen) from peripheral blood of MCI subjects and normal controls according to manufacturer’s instruction. Genotyping of NLRP3 SNPs was performed by melting curve genotyping method. This method consisted of a set of 3 primers including 2 allele-specific primers and a common reverse primer. The 2 allele-specific primers containing additional sequence of different length at 5′ end would produce polymerase chain reaction (PCR) products with different melting temperature due to differences in GC composition. A long 5′ GC sequence (5′-GCGGGCAGGGCGG-3′) and a short 5′ GC sequence (5′-GCGGGC-3′) was added to the allele-specific primers respectively. Genotypes at the SNP can be inferred from the different melting temperature of the PCR products. Melting curve analysis was performed by LC480 (Roche). Primers used in genotyping were listed in supplementary table 1.

### RNA extraction, reverse transcription and quantitative PCR (qPCR)

Total RNA was extracted from PBMC by Trizol LS and bromochloropropane (BCP), followed by isopropanol precipitation and 75% ethanol wash. miRNA was extracted from plasma by the same method with 1 μl glycogen (R0551, Thermo Fisher). Total RNA reverse transcription was performed by PrimeScript RT Reagent Kit (RR037A, Takara). Quantitative PCR was performed by TB Green Premix Ex Taq II (Tli RNase H Plus) kit (RR820A, Takara) in a volume of 10 μl, including 5 μl 2X SYBR Master Mix, 0.4 μl forward and reverse primer, 0.05 μl dUTP, 2.15 μl H2O and 2 μl cDNA. miRNA reverse transcription was performed by MicroRNA first-strand synthesis and miRNA quantitation kits (638315, TaKaRa). miRNA qPCR was performed in a volume of 10 μl, including 5ul 2X SYBR Master Mix, 0.4 μl forward and reverse primer, 3.2 μl H2O and 1 μl cDNA. Each sample was run in duplicate. The relative expression of target genes was normalized to UBC for mRNA and U6 for miRNA and calculated by 2 △△Ct method. qPCR was performed by LC480. Primers used in qPCR were listed in supplementary table 2.

### Plasmids

Gene fragments containing target SNPs were amplified by KAPA HiFi Hotstart ReadyMix PCR Kit (Roche). After purification, the PCR products were ligated to NheI digested pGL3-promoter vector by In-Fusion Cloning Kit (Takara) and transformed to Stbl3. Site-directed mutagenesis at selected SNPs was performed by KAPA HiFi. All purified plasmid sequences were confirmed by Sanger sequencing.

### Cell culture and transfection

HEK293FT cells were maintained in DMEM medium with 10% fetal bovine serum (FBS) and 1% penicillin/streptomycin in humidified incubator at 37C with 5% CO_2_. Transfection was performed using Lipofectamine 2000 when cells reached 50% confluency. To measure enhancer activity, 500ng pGL3-promoter vector and 50 ng pRL-CMV vector were transfected into HEK293FT cells for 48 hours. For miRNA interaction, 10nM miR-425-5p mimics or control mimics (Genepharma, China) were transfected into HEK293FT together with luciferase vectors.

### Luciferase reporter assay

Gene fragments of NLRP3 with wild type or mutant nucleotide was cloned into pGL3-promoter vector. 500 ng constructed pGL3-promoter vector or empty pGL3-promoter vector were cotransfected with 50 ng Renilla luciferase reporter vector pRL-CMV into HEK293FT cells. Dual-luciferase Reporter Assay kit (Promega) was used to measure firefly and renilla luciferase activity 48 hours after transfection. Relative luciferase activity was calculated as firefly luciferase activity divided by renilla luciferase activity. Each group was performed in triplicate.

### Statistical analysis

Deviations from the Hardy–Weinberg equilibrium (HWE) for genotypes at individual loci were assessed by using the Pearson chi-squared test with one degree of freedom. Comparison between two groups was carried out by Student’s t-test and three groups by one-way ANOVA with Bonferroni post hoc test. Chi-square test was used for genotype and allelic distribution of NLRP3 SNPs. Data were analyzed by GraphPad Prism 10.0 and SPSS 29.0. P-value<0.05 was considered as statistically significant.

## Results

### Association of NLRP3 SNPs with MCI

The genotype distribution of rs7525979 (p=0.046) and rs10754558 (p=0.049) were significantly different between MCI subjects and normal controls. Difference in allelic frequency was also found in rs7525979 (p=0.014). rs7525979 T allele carriers and rs10754558 C allele carriers were associated to significantly higher risk of developing MCI (Table 1). MoCA score less than 22 is usually considered as indication of cognitive impairment. When compared subjects with MoCA >=22 and subjects with MoCA < 22, rs3806268 was significantly different when compared minor allele genotype versus common allele genotype (Table 2).

**Table 1.**
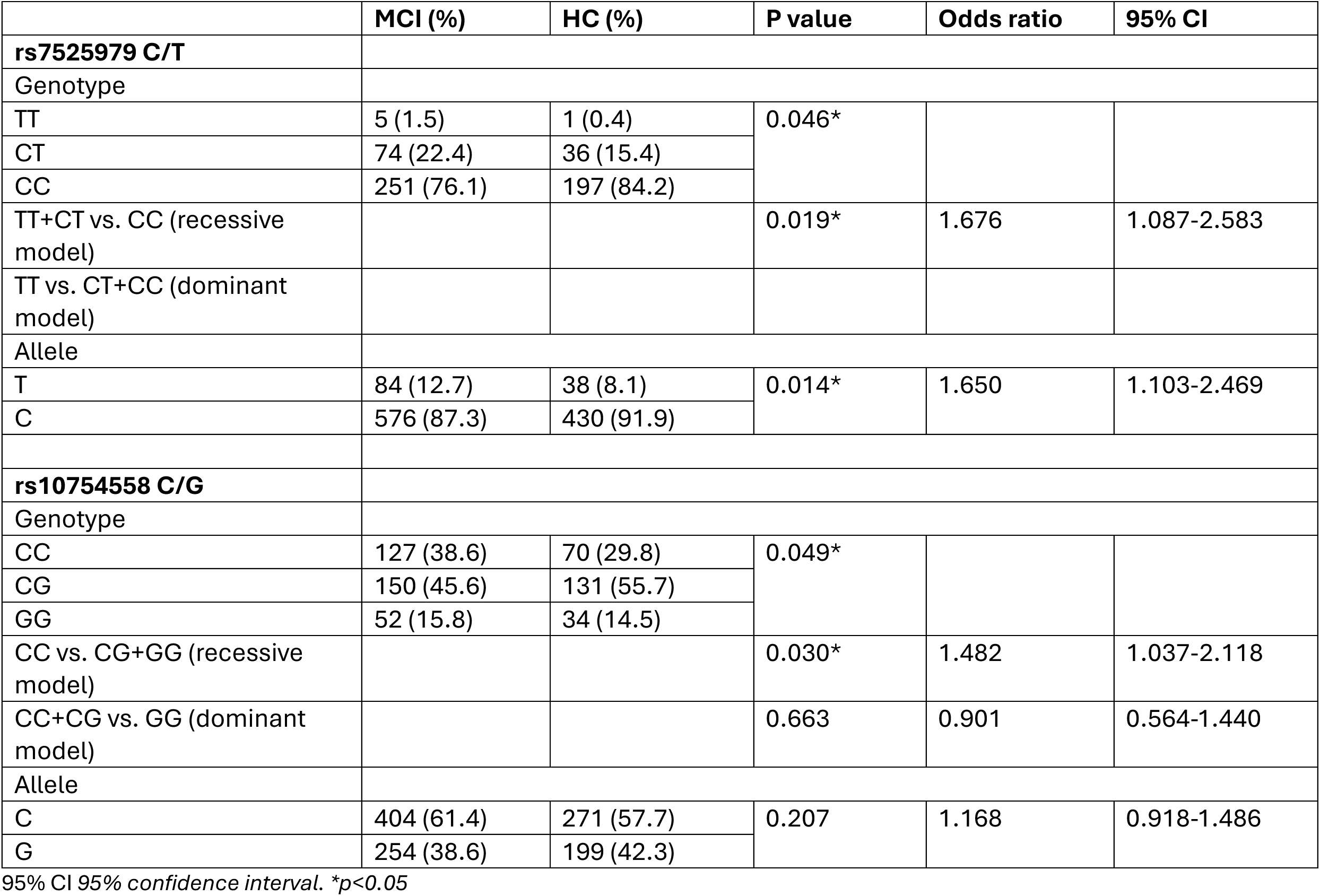
Genotype and allele distribution of NLRP3 SNPs in older adults with MCI and HC.

|  | MCI (%) | HC (%) | P value | Odds ratio | 95% CI |
| --- | --- | --- | --- | --- | --- |
| rs7525979 C/T |  |  |  |  |  |
| Genotype |  |  |  |  |  |
| TT | 5 (1.5) | 1 (0.4) | 0.046* |  |  |
| CT | 74 (22.4) | 36 (15.4) |  |  |  |
| CC | 251 (76.1) | 197 (84.2) |  |  |  |
| TT+CT vs. CC (recessive model) |  |  | 0.019* | 1.676 | 1.087-2.583 |
| TT vs. CT+CC (dominant model) |  |  |  |  |  |
| Allele |  |  |  |  |  |
| T | 84 (12.7) | 38 (8.1) | 0.014* | 1.650 | 1.103-2.469 |
| C | 576 (87.3) | 430 (91.9) |  |  |  |
| rs10754558 C/G |  |  |  |  |  |
| Genotype |  |  |  |  |  |
| CC | 127 (38.6) | 70 (29.8) | 0.049* |  |  |
| CG | 150 (45.6) | 131 (55.7) |  |  |  |
| GG | 52 (15.8) | 34 (14.5) |  |  |  |
| CC vs. CG+GG (recessive model) |  |  | 0.030* | 1.482 | 1.037-2.118 |
| CC+CG vs. GG (dominant model) |  |  | 0.663 | 0.901 | 0.564-1.440 |
| Allele |  |  |  |  |  |
| C | 404 (61.4) | 271 (57.7) | 0.207 | 1.168 | 0.918-1.486 |
| G | 254 (38.6) | 199 (42.3) |  |  |  |

95% CI 95% confidence interval. \* $p < 0.05$

**Table 2.** Genotype and allele distribution of NLRP3 SNPs in subjects with MoCA>=22 and MoCA<22.

| | MoCA≥22 (%) | MoCA$<$22 (%) | P value | Odds ratio | 95% CI |
| --- | --- | --- | --- | --- | --- |
| **rs3806268 A/G** |  |  |  |  |  |
| Genotype |  |  |  |  |  |
| AA | 83 (27.8) | 59 (36.6) | 0.138 |  |  |
| GA | 154 (51.5) | 71 (44.1) |  |  |  |
| GG | 62 (20.7) | 31 (19.3) |  |  |  |
| GG+GA vs. AA (recessive model) |  |  | 0.049* | 1.505 | 1.001 - 2.265 |
| GG vs. GA+AA (dominant model) |  |  | 0.706 | 1.097 | 0.678 - 1.775 |
| Allele |  |  |  |  |  |
| A | 320 (53.5) | 189 (58.7) | 0.131 | 1.235 | 0.939 - 1.624 |
| G | 278 (46.5) | 133 (41.3) |  |  |  |

95% CI 95% confidence interval. \* $p<0.05$

### Association between NLRP3 SNPs and inflammation related gene expression

There was positive correlation between NLRP3 mRNA level and scores of MoCA, but the level did not reach statistical significance. NLRP3 mRNA level was positively correlated with IL-1β, IL-18, IFN-γ, AIM2, CASP1, CASP5, PYCARD, TNF-α and NEK7 gene expression in PBMC (p<0.001) (Figure 2).

**Figure 2.**
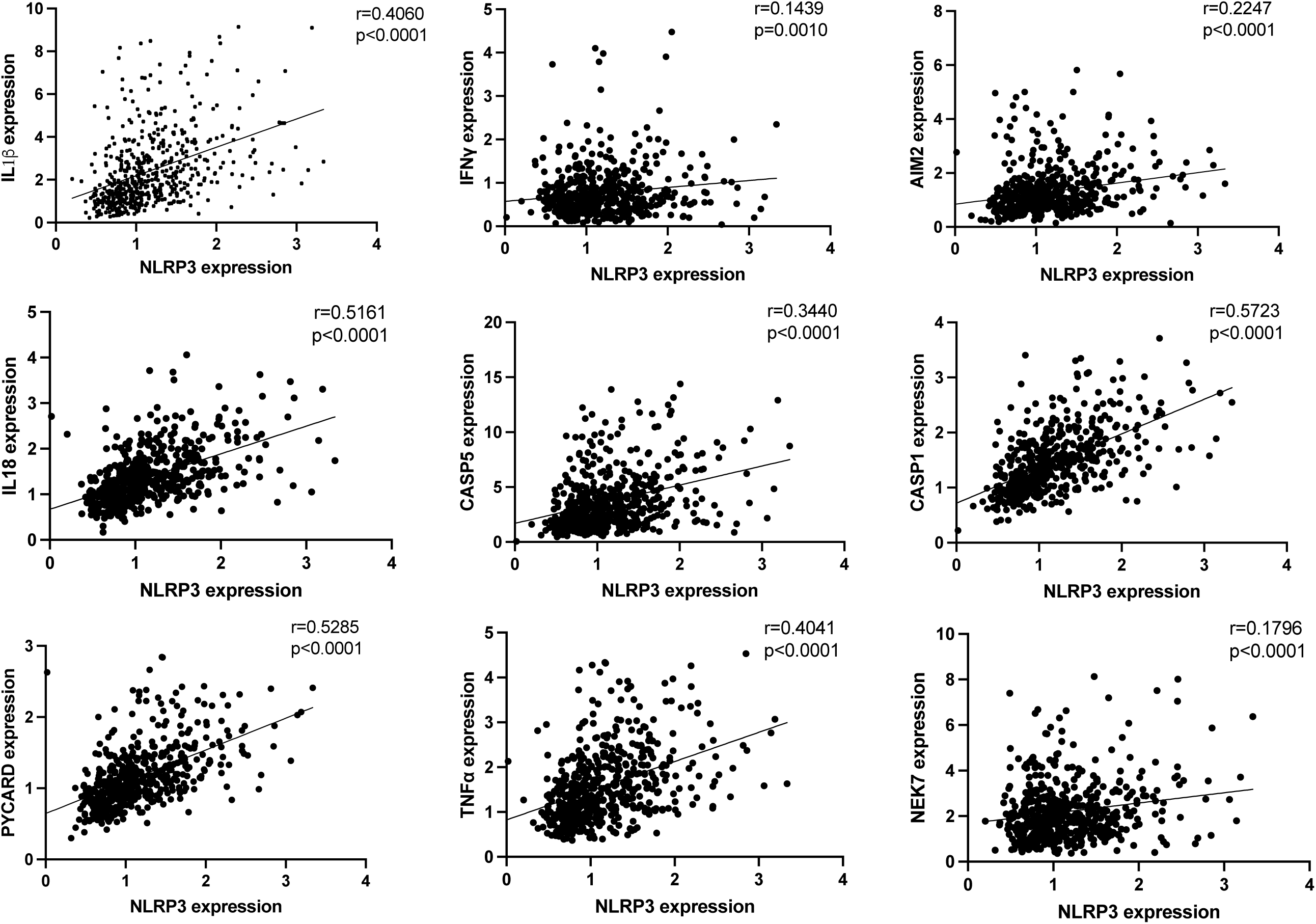
Correlation between NLRP3 and other inflammation related genes

The gene expression level of inflammatory related genes were quantified in MCI subjects and normal controls and the gene expression level was used to correlate with NLRP3 SNPs. rs12564791 was associated with the gene expression levels of NLRP3 (p=0.002), IL-18 (p=0.007), PYCARD (p=0.021) and CASP1 (p=0.023). rs10754557 was associated with the gene expression level of NEK7 (p=0.017) and GG/GA genotype was associated with the gene expression level of CASP5 (p=0.020). rs3806268 GG/GA genotype was associated with higher PYCARD gene expression (p=0.021). rs2027432 GG genotype was associated with higher PYCARD (p=0.021) and CASP1 (p=0.045) gene expression (Table 3, figure 3).

**Table 3.** (A) Association between NLRP3 SNPs genotype and inflammation related gene expression.

| SNP | NLRP3 | IL18 | PYCARD | CASP1 | CASP5 |
| --- | --- | --- | --- | --- | --- |
| **rs12564791** | 0.002* | 0.007* | 0.021* | 0.023* | 0.528 |
| **rs3806268** | 0.097 | 0.620 | 0.069 | 0.362 | 0.683 |
| **rs10754557** | 0.198 | 0.867 | 0.187 | 0.372 | 0.060 |

\*p<0.05

**Table 3.** (B) Association between NLRP3 SNPs recessive model and inflammation related gene expression.

| SNP | NLRP3 | IL18 | PYCARD | CASP1 | CASP5 |
| --- | --- | --- | --- | --- | --- |
| **rs12564791**<br>TT+CT vs. CC | 0.003* | 0.004* | 0.010* | 0.015* | 0.598 |
| **rs3806268**<br>GG+GA vs. AA | 0.088 | 0.345 | 0.021* | 0.175 | 0.414 |
| **rs10754557**<br>GG+GA vs. AA | 0.075 | 0.758 | 0.088 | 0.163 | 0.020* |
| **rs2027432**<br>AA+GA vs. GG | 0.159 | 0.272 | 0.039* | 0.045* | 0.419 |

\* $p < 0.05$

**Figure 3.**
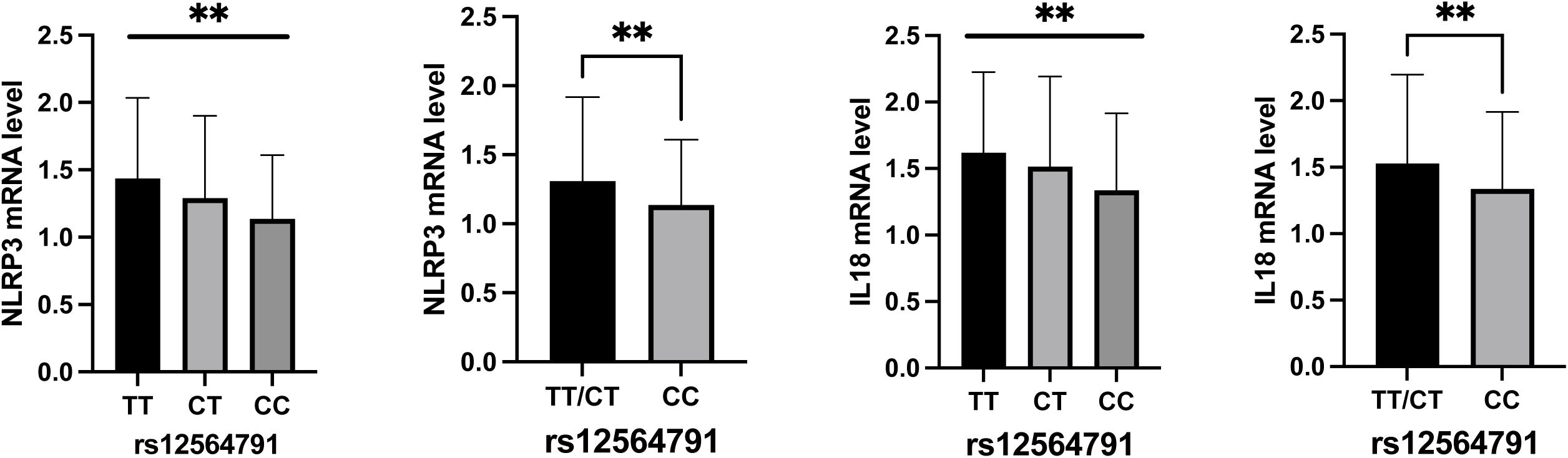
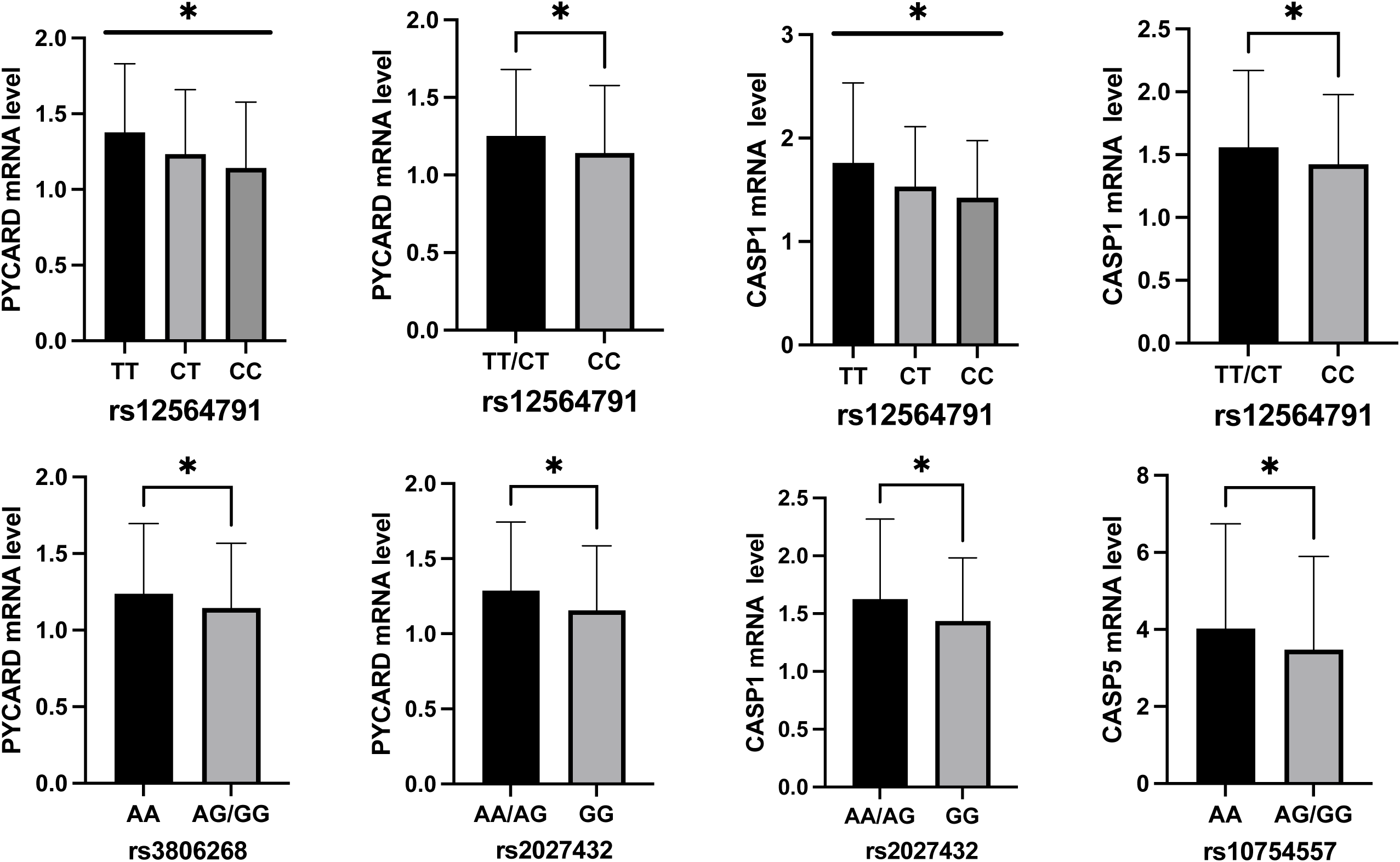
Association between NLRP3 SNPs and inflammation related gene expression. *\*p<0.05,**p<0.01*.

### Association of NLRP3 SNPs with cognitive decline

We next investigate whether NLRP3 SNPs were associated with cognitive decline. At 2-years follow-up, 15 out of 137 normal controls were progressed to MCI. Among those who were classified as MCI at baseline, 137 remains stable and two older adult was progressed to AD. rs7525979 CC/CT genotype, rs12048215 AA genotype and rs10754555 CC genotype were associated with MoCA score decrease>=2 at 2-years follow-up. Allelic distribution difference was also found in rs7525979 and rs10754555 (Table 4). The follow-up results showed that NLRP3 SNPs might have predictive value for cognitive decline.

**Table 4.** Association between NLRP3 SNPs with change in MoCA score.

|  | MoCA decrease>=2 (%) | MoCA decrease<2 (%) | P value | Odds ratio | 95% CI |
| --- | --- | --- | --- | --- | --- |
| rs10754555 C/G |  |  |  |  |  |
| Genotype |  |  |  |  |  |
| CC | 15 (50.0) | 43 (26.4) | 0.033* |  |  |
| GC | 13 (43.3) | 99 (60.7) |  |  |  |
| GG | 2 (6.7) | 21 (12.9) |  |  |  |
| GG+GC vs. CC (recessive model) |  |  | 0.010* | 2.791 | 1.259 - 6.186 |
| GG vs. GC+CC (dominant model) |  |  | 0.334 | 2.070 | 0.459 - 9.335 |
| Allele |  |  |  |  |  |
| C | 43 (71.7) | 185 (56.7) | 0.031* | 1.928 | 1.055 - 3.523 |
| G | 17 (28.3) | 141 (43.3) |  |  |  |
| rs12048215 A/G |  |  |  |  |  |
| Genotype |  |  |  |  |  |
| AA | 16 (53.3) | 55 (33.5) | 0.117 |  |  |
| GA | 12 (40.0) | 93 (56.7) |  |  |  |
| GG | 2 (6.7) | 16 (9.8) |  |  |  |
| GG+GA vs. AA (recessive model) |  |  | 0.038* | 0.442 | 0.201 - 0.970 |
| GG vs. GA+AA (dominant model) |  |  | 0.592 | 0.661 | 0.144 - 3.035 |
| Allele |  |  |  |  |  |
| A | 44 (73.3) | 203 (61.9) | 0.090 | 0.591 | 0.320 - 1.091 |
| G | 16 (26.7) | 125 (38.1) |  |  |  |
| rs7525979 C/T |  |  |  |  |  |
| Genotype |  |  |  |  |  |
| TT | 1 (3.4) | 1 (0.6) | 0.057 |  |  |
| CT | 10 (34.5) | 31 (19.0) |  |  |  |
| CC | 18 (62.1) | 131 (80.4) |  |  |  |
| CC vs. CT+TT (recessive model) |  |  | 0.029* | 2.521 | 1.084 - 5.861 |
| CC+CT vs. TT (dominant model) |  |  | 0.166 | 5.786 | 0.352 - 95.214 |
| Allele |  |  |  |  |  |
| T | 12 (20.7) | 33 (10.1) | 0.021* | 2.316 | 1.116 - 4.807 |
| C | 46 (79.3) | 293 (89.9) |  |  |  |

95% CI 95% confidence interval. \* $p < 0.05$

### rs10754555 affected enhancer activity of transcription

rs10754555 locates in the intronic region of NLRP3 and rs10754558 locates on 3’-UTR of NLRP3, we next investigate whether the G to C mutation of both SNPs enhance gene transcription. 300bp mRNA secondary structure was predicted by RNAfold Web Server (http://rna.tbi.univie.ac.at/cgi-bin/RNAWebSuite/RNAfold.cgi) and the result showed the RNA secondary structure containing C nucleotides of both SNPs was associated with a higher free energy and lower stability, when compared to the structure with G nucleotides. G-to-C mutation might change the structure of NLRP3 folding and therefore affected the downstream function. To confirm this prediction, pGL3-promoter vectors containing G/C were transfected into HEK293FT cells and luciferase reporter assay was performed. Vectors with G allele showed significantly higher luciferase activity, suggesting greater enhancer activity associated with G allele (Figure 4A-B).

**Figure 4.**
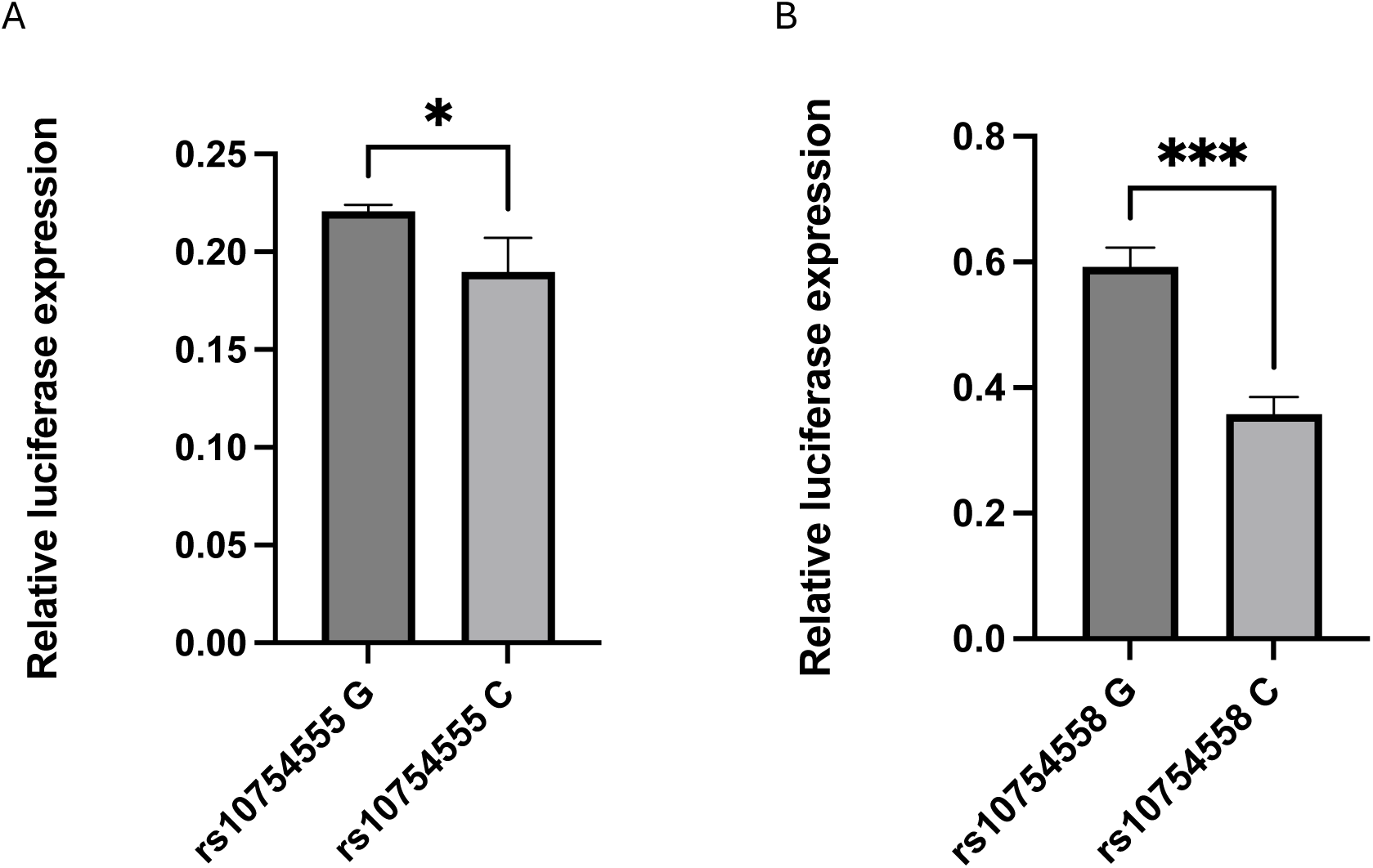
rs10754555 and rs10754556 affected enhancer activity. (A) transcriptional enhancer activities of rs10754555 G/C were detected using dual-luciferase reporter assay in HEK293FT cells; (B) transcriptional enhancer activities of rs10754558 G/C were detected using dual-luciferase reporter assay in HEK293FT cells. *\*p<0.05, ***p<0.001*.

### rs10754558 affected interaction with miR-425-5p

As rs10754558 locates within the 3’-UTR of NLRP3 gene, we further investigated whether it could affect the interactions with miRNAs. A SNP-miRNA interaction prediction tool, miRNASNP-v3(21), showed that rs10754558 is within the binding site of miR-425-5p and the G-to-C mutation disrupted this interaction (Figure 5A). To validate this interaction, pGL3-promoter vectors containing NLRP3 3’-UTR sequence with rs10754558 G or C were co-transfected with miR-425-5p mimics or negative control mimics into HEK293FT cells for 48 hours. The dual luciferase assay showed that miR-425-5p significantly reduced the luciferase activity in cells transfected with vector containing rs10754558 G allele. No obvious changes were found in cells transfected with rs10754558 C allele (Figure 5B). This result indicated that NLRP3 is a target gene of miR-425-5p and rs10754558 G-to-C mutation affected the binding of miR-425-5p to NLRP3 3’-UTR.

**Figure 5.**
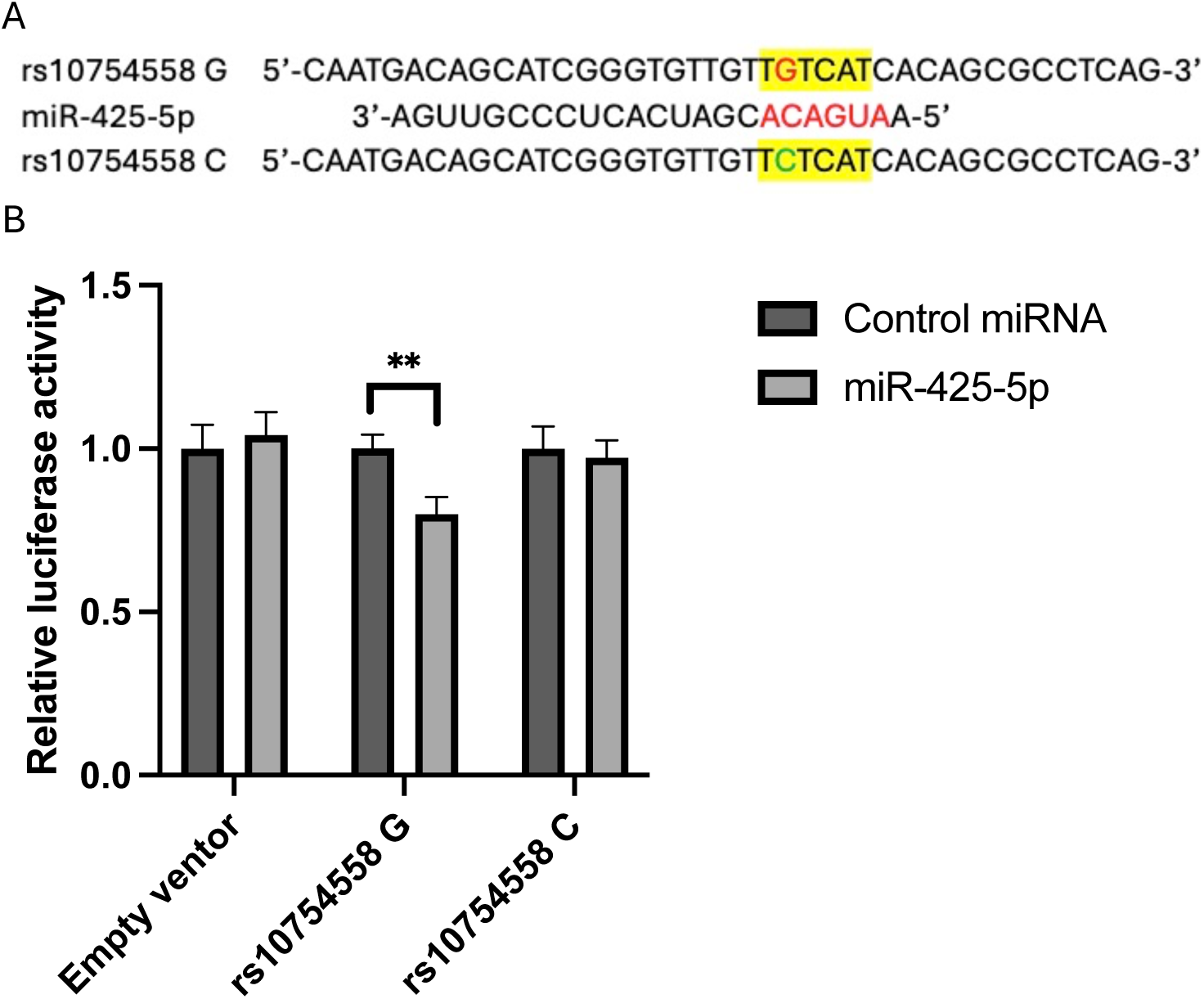
rs10754558 affects interaction with miR-425-5p. (A) rs10754558 G to C mutation was predicted to disturb the binding between NLRP3 3’-UTR and miR-425-5p; (B) The binding activities of rs10754558 G/C with miR-425-5p were determined by dual-luciferase reporter assay in HEK293FT cells. *\*\*p<0.01*.

### miR-425-5p negatively correlated with MoCA

Plasma RNA was extracted from 26 MCI subjects and 26 age and gender matched normal controls and reverse transcription was performed by adding Poly(A) tail to the 3’ end of miRNA. The results showed that plasma miR-425-5p expression was negative correlated with MoCA score (p=0.033) (Figure 6A). When using the median of miR-425-5p expression level to define high and low expression groups, MoCA score were significantly different between the two groups. High miR-425-5p expression group was associated to lower average MoCA score (p=0.011) (Figure 6B).

**Figure 6.**
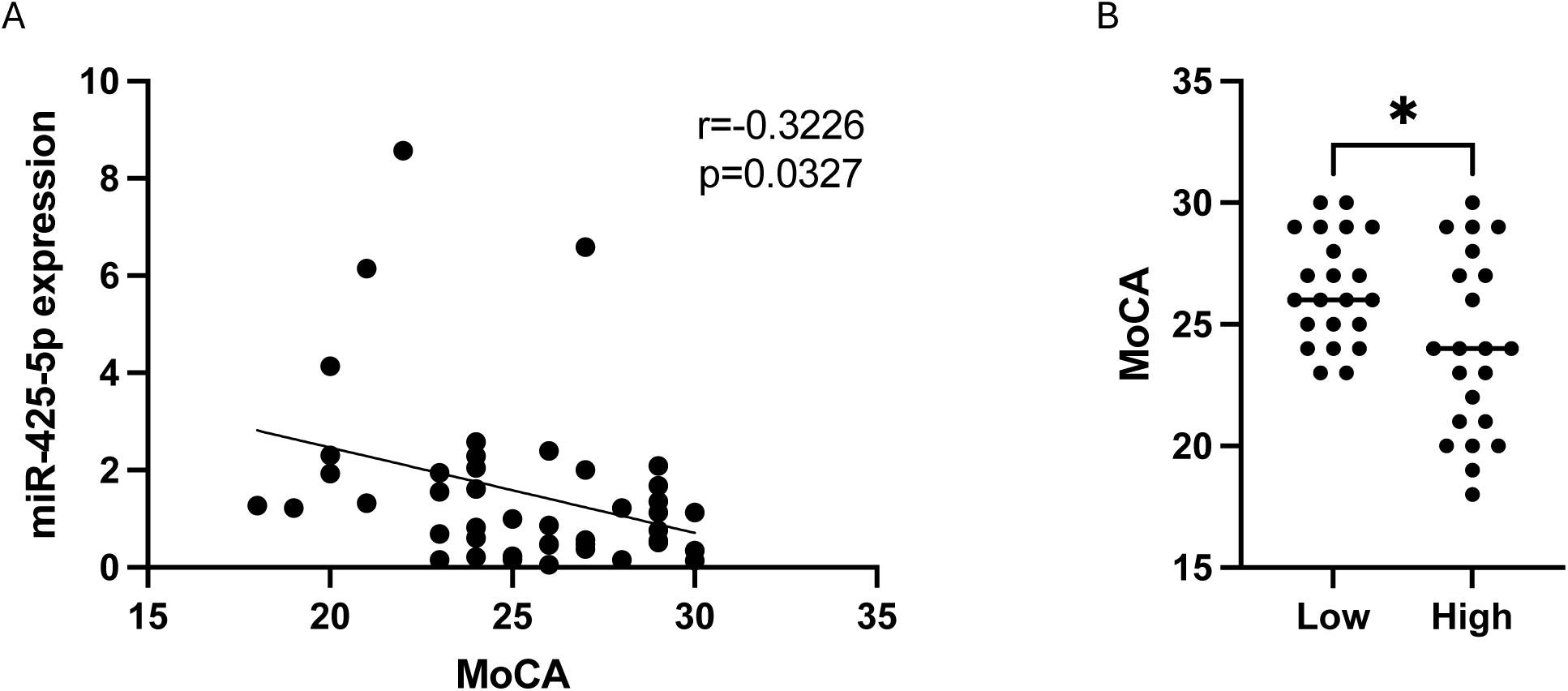
miR-425-5p negatively correlateed with MoCA. (A) miR-425-5p expression negatively correlateed with MoCA score; (B) MoCA score is different between miR-425-5p expression high or low group. *\*p<0.05*.

## Discussion

Increasing evidence demonstrate the important role of inflammation in MCI and AD pathogenesis. NLRP3 is a key inflammasome participating in inflammation and inflammatory cytokines production. Genetic variations of NLRP3 have been identified in multiple inflammatory related disease, including inflammatory bowel disease, periodontitis, asthma, primary gouty arthritis and autoimmune thyroid disease(22–26). Previous studies have found that rs2027432 was associated with late onset AD (LOAD) risk and rs10754558 was associated with LOAD only in ApoE ε4 carriers(18). Our study identified the associations between NLRP3 SNPs rs7525979 and rs10754558 and the onset of MCI in Chinese. Individuals carrying at least one T allele of rs7525979 or one C allele of rs10754558 had higher risk of developing MCI than those without copies of the risk allele. In addition, rs10754558 could affect NLRP3 mRNA level and increased mRNA stability(11,13). rs10754558 G to C mutation was demonstrated to alter the interaction between NLRP3 and miR-146a-5p, which further confirmed this SNP may be functional(12). Our study also showed that rs10754558 might regulate NLRP3 gene expression and affect the interaction with miR-425-5p. rs7525979, a synonymous SNP located in exon 3, have been shown to affect the efficiency of NLRP3 translation, impact NLRP3 protein stability, ubiquitination state, and solubility(27). Synonymous mutations leading to rare codons were usually considered to be translated more slowly(28). The deregulated translation rate may cause protein misfolding, which was widely recognized to affect proteinopathies like AD(27). In our finding, rs7525979 was associated with MCI risk and cognitive decline. Another synonymous variant rs3806268 was associated with the gene expression of PYCARD. The result might be explained by the altered translation efficacy and protein folding. This in turn influenced protein function and downstream pathways which further result in disease progression. NLRP3 SNPs were also associated with cognitive decline during a 2-year follow up. rs12048215 and rs10754555 were associated with decline in MoCA score. rs10754555 G to C mutation affected transcriptional activity, which may result in altered gene expression and NLRP3 function and therefore influence cognition.

Although introns consist of 90% of gene sequence, most functional mutations are found in exons(29). Recent studies have highlighted the impact of intron variants to regulate splicing efficiency and abnormal splicing including exon skipping and intron inclusion(29,30). Intronic SNPs could also influence allele imprinting, gene expression and cell apoptosis(31). Most of the SNPs investigated in our study are located in intron regions. rs12564791 was associated with the gene expression level of NLRP3 and IL-18. The expression of PYCARD and CASP1 were associated with rs12564791and rs2027432. rs10754557 was associated with the gene expression level of NEK7 and CASP5. NLRP3 consists of the sensor NLRP3, the adaptor PYCARD and the effector pro-caspase1. Upon inflammasome activation, NLRP3 recruits PYCARD and forms a large filament molecule ASC speck. Assembled ASC speck recruits pro-caspase 1 and enables caspase 1 self-cleavage and activation. At the meantime, the interaction between NEK7 and NLRP3 increases which is essential for ASC speck formation and caspase 1 activation(1). Caspase1 further cleaves the cytokine precursors, pro-IL-18 and pro-IL-1β, to their active form IL-18 and IL-1β. The intron SNPs of NLRP3 affected gene expression of NLRP3 inflammasome components and also downstream cytokines, indicating their potential role in modulating inflammasome assembling and function.

Previous study showed gene expression level of NLRP3 but no other related molecules was upregulated in stimulated monocytes of MCI subjects(2). However, no significant difference of any inflammation gene expression between MCI subjects and normal controls was found in our study. This may be due to different experimental procedures as our study assessed mRNA level directly in PBMC. In the initial stage of the disease, inflammation genes may start to express abnormally but not yet reach obvious changes without extra stimulations. The expression of NLRP3 was positively correlated with all other inflammation genes, indicating the potential assembly step as the disease progress.

miRNAs are small, single-stranded, non-coding RNA that play crucial roles in regulating gene expression, as well as cell proliferation, apoptosis, invasion and migration. miRNAs have been implicated in AD pathogenesis and the expression of several miRNAs are reported to be different between AD patients and healthy individuals in brain, cerebrospinal fluid, and blood(32). miRNAs achieve their function by binding to the 3’-UTR of target genes and further lead to the upregulation or downregulation of target genes. Therefore, mutations within 3’-UTR may affect the interactions between target genes and miRNAs. rs10754558 G to C mutation was predicted to be a binding site of miR-425-5p. Our functional study confirmed that the 3’UTR SNP rs10754558 might affect the interaction with miR-425-5p. miR-425-5p has been shown to be upregulated in plasma of AD patients, which might be a potential biomarker(33), miR-425-5p may participate in AD by interacting with BACE1 and HSPB8 and further regulate BACE1 activity, promote tau phosphorylation and cell apoptosis (33,34). miR-425-5p was negatively associated with blood-based inflammation markers C-reactive protein and fibrinogen, indicating its role in inflammation(35). Our results showed that plasma miR-425-5p expression was negatively correlated with MoCA score, which was in line with previous study. However, our result did not show any difference between MCI subjects and normal controls regarding miR-425-5p expression level, probably due to the small sample size.

In summary, our study suggested that NLRP3 SNPs might be associated with cognitive decline in Chinese. There are a few limitations in the present study. This project only included Chinese Han population with a small sample size. Future research in other diverse populations and longer follow up might be needed to confirm the finding.

## Data availability

All data generated or analyzed during this study are included in this article. Further enquiries can be directed to the corresponding author.

## Ethical approval

The study was approved by the Clinical Research Ethics Committee of the Chinese University of Hong Kong.

## Consent for publication

Not applicable.

## Acknowledgement

The study was supported by the Health and Medical Research Fund (Ref no. 09200246), Health Bureau, Hong Kong SAR Government and Direct Grant from the Chinese University of Hong Kong (Ref no. 2021.057). The authors appreciate the support from Dr. Raymond Wai Ming Lung and Dr. Chit Chow.

## Author’s contributions

R.G was responsible for conducting experiments, data analysis and preparing manuscript ; C.W.L was responsible for clinical supervision; S.L.M was responsible for manuscript revision. All authors contributed to reviewing the manuscript and approving the final version.

## References

1. He Y, Zeng MY, Yang D, Motro B, Núñez G. NEK7 is an essential mediator of NLRP3 activation downstream of potassium efflux. Nature. 2016 Feb;530(7590):354–7.

2. Saresella M, La Rosa F, Piancone F, Zoppis M, Marventano I, Calabrese E, et al. The NLRP3 and NLRP1 inflammasomes are activated in Alzheimer’s disease. Mol Neurodegeneration. 2016 Dec;11(1):23.

3. Ahmed ME, Iyer S, Thangavel R, Kempuraj D, Selvakumar GP, Raikwar SP, et al. Co-Localization of Glia Maturation Factor with NLRP3 Inflammasome and Autophagosome Markers in Human Alzheimer’s Disease Brain. JAD. 2017 Oct 3;60(3):1143–60.

4. Heneka MT, Kummer MP, Stutz A, Delekate A, Schwartz S, Vieira-Saecker A, et al. NLRP3 is activated in Alzheimer’s disease and contributes to pathology in APP/PS1 mice. Nature. 2013 Jan;493(7434):674–8.

5. Dempsey C, Rubio Araiz A, Bryson KJ, Finucane O, Larkin C, Mills EL, et al. Inhibiting the NLRP3 inflammasome with MCC950 promotes non-phlogistic clearance of amyloid-β and cognitive function in APP/PS1 mice. Brain, Behavior, and Immunity. 2017 Mar;61:306–16.

6. Jayaraman A, Htike TT, James R, Picon C, Reynolds R. TNF--mediated neuroinflammation is linked to neuronal necroptosis in Alzheimer’s disease hippocampus. acta neuropathol commun. 2021 Dec;9(1):159.

7. Cheng JJ, Ma XD, Ai GX, Yu QX, Chen XY, Yan F, et al. Palmatine Protects Against MSU-Induced Gouty Arthritis via Regulating the NF-κB/NLRP3 and Nrf2 Pathways. DDDT. 2022 Jul;Volume 16:2119–32.

8. Meimei C, Fei Z, Wen X, Huangwei L, Zhenqiang H, Rongjun Y, et al. Taxus chinensis (Pilg.) Rehder fruit attenuates aging behaviors and neuroinflammation by inhibiting microglia activation via TLR4/NF-κB/NLRP3 pathway. Journal of Ethnopharmacology. 2025 Jan;337:118943.

9. Bate C, Kempster S, Last V, Williams A. Interferon-γ increases neuronal death in response to amyloid-β1-42. J Neuroinflammation. 2006;3(1):7.

10. Kaabi YA. The NLRP3 inflammasome rs35829419 C>A polymorphism is associated with type 2 diabetes mellitus in Saudi Arabia. SMJ. 2023 Aug;44(8):745–50.

11. Hitomi Y, Ebisawa M, Tomikawa M, Imai T, Komata T, Hirota T, et al. Associations of functional NLRP3 polymorphisms with susceptibility to food-induced anaphylaxis and aspirin-induced asthma. Journal of Allergy and Clinical Immunology. 2009 Oct;124(4):779–785.e6.

12. Lu F, Chen H, Hong Y, Lin Y, Liu L, Wei N, et al. A gain-of-function NLRP3 3′-UTR polymorphism causes miR-146a-mediated suppression of NLRP3 expression and confers protection against sepsis progression. Sci Rep. 2021 Jun 25;11(1):13300.

13. Zhu Z, Yan J, Geng C, Wang D, Li C, Feng S, et al. A Polymorphism Within the 3′UTR of NLRP3 is Associated with Susceptibility for Ischemic Stroke in Chinese Population. Cell Mol Neurobiol. 2016 Aug;36(6):981–8.

14. Roberts RL, Van Rij AM, Phillips LV, Young S, McCormick SPA, Merriman TR, et al. Interaction of the inflammasome genes CARD8 and NLRP3 in abdominal aortic aneurysms. Atherosclerosis. 2011 Sep;218(1):123–6.

15. Zhang AQ, Zeng L, Gu W, Zhang LY, Zhou J, Jiang D po, et al. Clinical relevance of single nucleotide polymorphisms within the entire NLRP3 gene in patients with major blunt trauma. Crit Care. 2011;15(6):R280.

16. Awni AA, Hamed ZO, Abdul-Hassan Abbas A, Abdulamir AS. Effect of NLRP3 inflammasome genes polymorphism on disease susceptibility and response to TNF--α inhibitors in Iraqi patients with rheumatoid arthritis. Heliyon. 2023 Jun;9(6):e16814.

17. Kumar N, Kaur M, Singh G, Valecha S, Khinda R, Di Napoli M, et al. A susceptibility putative haplotype within NLRP3 inflammasome gene influences ischaemic stroke risk in the population of Punjab, India. Int J Immunogenetics. 2022 Aug;49(4):260–70.

18. Tan MS, Yu JT, Jiang T, Zhu XC, Wang HF, Zhang W, et al. NLRP3 polymorphisms are associated with late-onset Alzheimer’s disease in Han Chinese. Journal of Neuroimmunology. 2013 Dec;265(1–2):91–5.

19. Wong A, Xiong YY, Kwan PWL, Chan AYY, Lam WWM, Wang K, et al. The Validity, Reliability and Clinical Utility of the Hong Kong Montreal Cognitive Assessment (HK-MoCA) in Patients with Cerebral Small Vessel Disease. Dement Geriatr Cogn Disord. 2009;28(1):81–7.

20. †The International HapMap Consortium. The International HapMap Project. Nature. 2003 Dec;426(6968):789–96.

21. Liu CJ, Fu X, Xia M, Zhang Q, Gu Z, Guo AY. miRNASNP-v3: a comprehensive database for SNPs and disease-related variations in miRNAs and miRNA targets. Nucleic Acids Research. 2021 Jan 8;49(D1):D1276–81.

22. Heidari Z, Salimi S, Rokni M, Rezaei M, Khalafi N, Shahroudi MJ, et al. Association of IL-1β, NLRP3, and COX-2 Gene Polymorphisms with Autoimmune Thyroid Disease Risk and Clinical Features in the Iranian Population. Biomed Res Int. 2021;2021:7729238.

23. de Alencar JB, Zacarias JMV, Tsuneto PY, Souza VH de, Silva C de OE, Visentainer JEL, et al. Influence of inflammasome NLRP3, and IL1B and IL2 gene polymorphisms in periodontitis susceptibility. PLoS One. 2020;15(1):e0227905.

24. Queiroz G de A, da Silva RR, Pires A de O, Costa RDS, Alcântara-Neves NM, da Silva TM, et al. New variants in NLRP3 inflammasome genes increase risk for asthma and Blomia tropicalis-induced allergy in a Brazilian population. Cytokine X. 2020 Sep;2(3):100032.

25. Meng DM, Zhou YJ, Wang L, Ren W, Cui LL, Han L, et al. Polymorphisms in the NLRP3 gene and risk of primary gouty arthritis. Mol Med Rep. 2013 Jun;7(6):1761–6.

26. Lazaridis LD, Pistiki A, Giamarellos-Bourboulis EJ, Georgitsi M, Damoraki G, Polymeros D, et al. Activation of NLRP3 Inflammasome in Inflammatory Bowel Disease: Differences Between Crohn’s Disease and Ulcerative Colitis. Dig Dis Sci. 2017 Sep;62(9):2348–56.

27. von Herrmann KM, Salas LA, Martinez EM, Young AL, Howard JM, Feldman MS, et al. NLRP3 expression in mesencephalic neurons and characterization of a rare NLRP3 polymorphism associated with decreased risk of Parkinson’s disease. NPJ Parkinsons Dis. 2018;4:24.

28. Chaney JL, Clark PL. Roles for Synonymous Codon Usage in Protein Biogenesis. Annu Rev Biophys. 2015 Jun 22;44(1):143–66.

29. Seo S, Takayama K, Uno K, Ohi K, Hashimoto R, Nishizawa D, et al. Functional Analysis of Deep Intronic SNP rs13438494 in Intron 24 of PCLO Gene. Liu C, editor. PLoS ONE. 2013 Oct 22;8(10):e76960.

30. Chiang HL, Wu JY, Chen YT. Identification of functional single nucleotide polymorphisms in the branchpoint site. Hum Genomics. 2017 Dec;11(1):27.

31. Maan Hasan Salih, Adnan F Al-Azzawie, Akeel Hussain Ali Al-Assie. Intronic SNPs and Genetic Diseases: A Review. Int j res appl sci biotechnol. 2021 Apr 20;8(2):267–74.

32. Takousis P, Sadlon A, Schulz J, Wohlers I, Dobricic V, Middleton L, et al. Differential expression of microRNAs in Alzheimer’s disease brain, blood, and cerebrospinal fluid. Alzheimer’s Camp; Dementia. 2019 Nov;15(11):1468–77.

33. Yuan J, Wu Y, Li L, Liu C. MicroRNA-425-5p promotes tau phosphorylation and cell apoptosis in Alzheimer’s disease by targeting heat shock protein B8. J Neural Transm. 2020 Mar;127(3):339–46.

34. Samadian M, Gholipour M, Hajiesmaeili M, Taheri M, Ghafouri-Fard S. The Eminent Role of microRNAs in the Pathogenesis of Alzheimer’s Disease. Front Aging Neurosci. 2021 Mar 15;13:641080.

35. Van der Auwera S, Ameling S, Wittfeld K, Frenzel S, Bülow R, Nauck M, et al. Circulating microRNA miR-425-5p Associated with Brain White Matter Lesions and Inflammatory Processes. Int J Mol Sci. 2024 Jan 10;25(2):887.

